# High Intensity Focused Ultrasound (HIFU) Thalamotomy as a Safe and Effective Treatment of Essential Tremor (ET) in Older Populations and those with Comorbidities

**DOI:** 10.64898/2026.10.14.26365548

**Authors:** Adam Farmer, William Young, Jody Neer, Fen-Lei Chang, Tara Kimbason

## Abstract

Magnetic Resonance guided High Intensity Focused Ultrasound (MRgHIFU) has been demonstrated to be a safe and effective minimally invasive therapy for essential tremor (ET). However, it is not clear whether older adults (≥70 years old) or individuals with comorbidities exhibit similar safety and efficacy. This IRB approved project collected EPIC EMR data on patients that underwent MRgHIFU at Parkview Regional Medical Center from January 2023 to November 2025. 68 patients were assessed for tremor severity using Clinical Rating Scale for Tremor (CRST), quality of life using Quality of life in ET Questionnaire (QUEST), and safety profile via reported post-procedure side effects. CRST and QUEST scores improved up to 1 year from baseline in our patient cohort and efficacies in older (≥70 years of age) and younger (<70 years of age) patients up to 3 months post MRgHIFU were not statistically different. Effects of comorbidities on CRST or QUEST scores improvements were all transient or did not affect endpoints. Older patients showed fewer total side effects at 3 months (1.17 (95% CI: 0.47-1.87) vs 0.21 (95% CI: −0.09-0.51), p=0.017) as did patients with pulmonary disease at 1 month (1.9 (95% CI: 1.6-2.2) vs 1.1 (95% CI: 0.9-1.3), p=0.036) while patients with family history of tremor showed more side effects at day 1 (1.5 (95% CI: 0.8-2.2) vs 2.5 (95% CI: 2.1-2.9), p=0.025). Age and comorbidities were not detected to adversely affect safety or efficacy following MRgHIFU in ET patients. Further studies with larger cohorts are needed for validation of findings and assessment for equivalence.

## Introduction

Essential tremor (ET) is one of the most common movement disorders in both the US and worldwide. Global prevalence of ET is between 0.67% and 1.33% and the prevalence in the US is 2.2% skewed towards older individuals (1–2). ET is defined as being bilateral upper extremity action tremor with at least a three-year duration (3–4). However, the tremor can also affect the head, voice, legs, face, trunk and tongue at a lower frequency (5). ET can greatly diminish the quality of life of those affected and some data suggests that individuals with ET have an overall higher mortality rate than the general population (6–7). ET can be a source of both social embarrassment and disability in the lives of individuals, often leading to earlier retirement (6, 8).

The underlying pathology of ET has been shown to involve both indirect and direct cerebellar dysfunction with indirect causes due to locus coeruleus dysfunction and direct causes due to Purkinje cell dysfunction (9). However, the precise order of the pathophysiological events and the underlying causes are unknown. While ET has a strong genetic influence with 30-70% of patients with ET having a family history of typically autosomal dominant inheritance, the underlying genetic causes are not well established though many candidate genes have been identified with LINGO1 and STK32B (5, 10).

The severity of ET in patients can be graded by two validated assessments known as The Quality of Life in ET Questionnaire (QUEST) and Clinical Rating Scale for Tremor (CRST) to assess progression or alleviation of ET (11–12). QUEST is a 30-part questionnaire used to measure quality of life in patients with ET while CRST is an evaluation with three subcategories that can be further analyzed to evaluate the physical severity of ET (11, 13). Part A is tremor severity as ranked by clinician, part B is motor tasks as ranked by clinician, and part C is functional disability as ranked by patient. Together these two scores can be used to monitor treatment effectiveness of ET.

Individuals learn to adapt and live with tremors, but pharmacological treatments are considered as ET progresses. The primary first line pharmacological treatments are propranolol or primidone, but multiple second line treatments are also available (14). Given that up to 50% of patients with ET are either refractory to or cannot tolerate pharmaceutical treatment, there is a need for alternative interventional therapies such as Magnetic Resonance guided High Intensity Focused Ultrasound (MRgHIFU) (15–16). MRgHIFU has demonstrated in recent years to be a safe and effective minimally invasive therapy for ET (15, 17). Compared to DBS, MRgHIFU has a similar efficacy and safety profile with the advantage of being less invasive (18–21). MRgHIFU treats ET by lesioning the ventral intermediate (Vim) nucleus of the thalamus using heat generated from focused ultrasound waves thereby disrupting neural pathways that facilitate ET (22). The ideal location for lesioning has recently been further defined at roughly 1.2-1.5 mm above the anterior commissure-posterior commissure (AC-PC) axis in the Vim nucleus of the thalamus (23). The sustained benefit of MRgHIFU in the treatment of ET have recently been shown to last up to five years following the intervention suggesting that the effects are long lasting (24–25). While MRgHIFU has been shown to be effective at treating ET and improving the quality of life of those affected, it is not clear what factors predict safety and efficacy.

A previous study recently published showed that adults older than 70 years old showed a similar efficacy and safety profile when examining side effects and CRST scores (26). However, neither QUEST scores or comorbidities outside of vascular pathologies were assessed, giving an incomplete comparative picture of how ET treatment affects quality of life of older adults and those with comorbidities. This is especially critical to understand as ET prevalence and comorbidities increases with age. Additionally, it is not known whether factors such as family history of tremor or concurrent neurological conditions (CNCs) predict outcomes. Finally, the only comorbidities that have been investigated as possibly contributing to MRgHIFU outcomes for essential tremor are vascular in origin (26).

The objectives of this study are to understand if and how age and common comorbidities affect efficacy and safety of MRgHIFU thalamotomy in patients with ET.

## Materials and Methods

### Data Collection

This IRB approved retrospective chart review project collected EPIC EMR data on patients that underwent MRgHIFU at Parkview Regional Medical Center under the direction of Neurosurgery and Neurology from January 2023 to November 2025. Inclusion criteria included individuals with clinical diagnosis of ET and ≥ 18 years old. Exclusion criteria included patients who did not have a consultation visit confirming essential tremor diagnosis, patients <18 years of age, and patients not meeting the inclusion criteria. Time points collected were baseline, 1 day, 1 week, 1 month, 3 months, and 1 year. Sixty-eight patients were analyzed for changes in CRST and specific subcategories to assess tremor severity and QUEST to assess life quality following treatment scores before and after MRgHIFU to assess efficacy. CRST scores were further broken down and analyzed for upper extremity specific scores by totaling Subcategory A and Subcategory B (Sub A+Sub B) scores specific to the treated side and analysis of Subcategory C (Sub C) scores that indicate functional disability experienced by the patient due to their tremor. Total side effects were quantified based on categories of side effects classified in EPIC.

Categories of side effects included paresthesia/numbness/tingling, changes to taste or smell, tongue biting, hiccups, speech changes, dysmetria, gait/balance changes, sleep changes, headaches >24 hours, and weakness. Additionally, comorbidities, family history, and demographic data (age, sex, race, BMI) were collected from EPIC for further analysis as shown in Table 1. All subgroup analyses (by age, comorbidity, and family history) are exploratory and hypothesis-generating in nature.

**Table 1.**
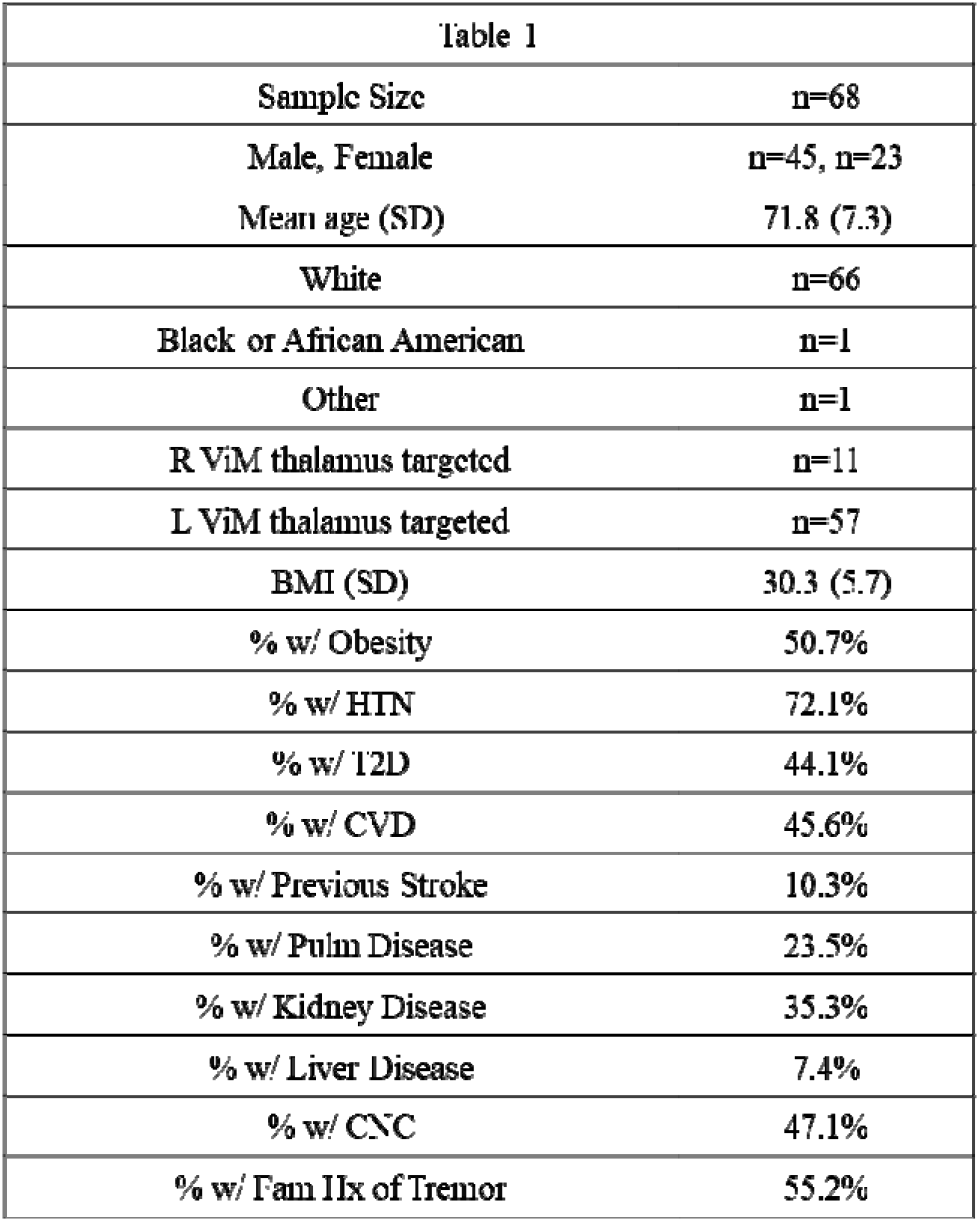
Data collected from EPIC records for the 68 patients in this study was quantitated and shown in table. Note that age, comorbidities, and family history (Fam Hx) were assessed for being possible predictors of efficacy and safety. BMI could not be calculated for one patient due to missing height in EMR. Note that the most common CNCs were Parkinson disease (n=10), neuropathy (n=10), and cognitive dysfunction (n=8). SD=standard deviation; HTN=hypertension; T2D=type II diabetes; Pulm Dis=pulmonary disease; CNC=concurrent neurological condition; L=left; Vim=ventral intermediate; R=right.

### Data Analysis

Statistical tests performed included two tailed independent means t-tests assuming equal or unequal variances based on F test results were used to examine differences in efficacy and safety between older adults ≥70 years of age and <70 years of age as well as individuals with comorbidities or family history of ET. If differences in efficacy as shown by improvements in CRST or QUEST scores were seen, baseline and time points of interest were compared for both differences in starting in end point data that could affect improvement data. Paired two tailed t-tests were used to assess efficacy of MRgHIFU on ET at different time points for the same patients. All stats were calculated in Microsoft Excel and evaluated based on a 0.05 significance level. Confidence intervals were based on estimated confidence intervals (CI) using CONFIDENCE.T function in excel. Lesioning parameters (device, skull density ratio, sonication number, max temperature, lesion size) were not systematically analyzed or collected. No formal correction for normality or multiple comparisons was performed given excel institutional constraints and data privacy.

## Results

### Characteristics of patient population

Demographic data pooled from EPIC for those who underwent MRgHIFU were examined to assess patient population characteristics (Table 1). Overall, the patient population was predominantly Caucasian men with average age of 72 years, obese, and with underlying hypertension.. Additionally, the left ViM thalamus was the most common targeted side as the majority of the patients treated are right-handed.

### MRgHIFU is effective at improving CRST scores up to 1 year post HIFU

Previous findings have demonstrated that ET treatment from MRgHIFU can last up to five years (24–25). To assess whether long term results up to 1 year were found in our cohort, changes in both total CRST, Sub A+Sub B treated hand specific CRST sum, and CRST Sub C scores were assessed. Total CRST scores give an overall view of bilateral ET severity in general while the Sub A+B treated hand scores give a more precise view of tremor severity and its impact on tasks on the affected side. Finally, Sub C scores assess the extent of functional disability the patient experiences as a result of their tremor.

Overall, all CRST scores assessed were found to be reduced up to 1 year post HIFU for the treated cohort (Figure 1A-C). These findings suggest that MRgHIFU leads to long term benefits in our treated cohort for both treated hand specific tremor severity as well as the functional disabilities experienced by patients with ET.

**Figure 1.**
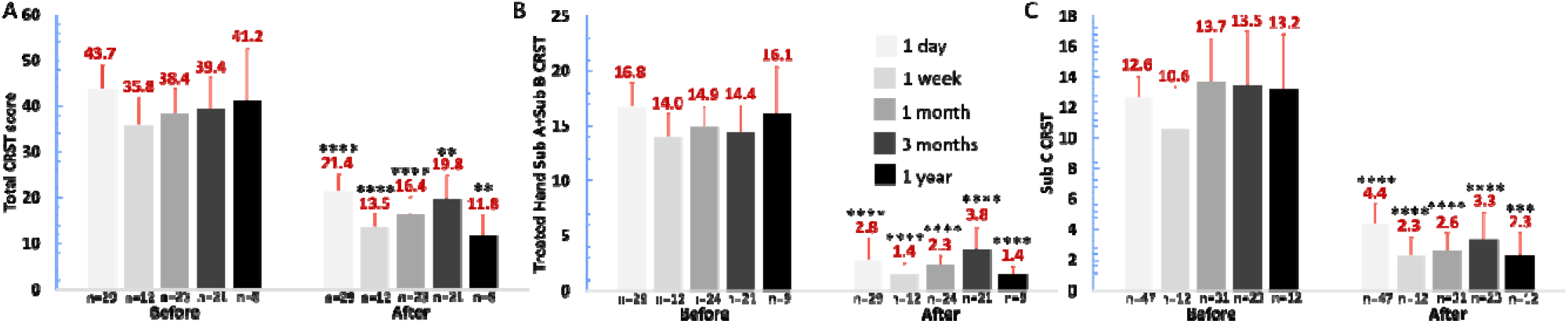
Total and CRST Sub scores improve up to 1-year post-HIFU. A: Total CRST scores over time post-HIFU. Total CRST scores show a statistically significant decrease at all points measured. B: Treated Hand Sub A+Sub B scores over time post-HIFU More precise measurements of tremor severity in the affected upper extremity demonstrate consistent decreases in tremor severity at each time point measured. C: Sub C CRST score improvements. Sub\ C scores measuring functional disability related to ET showed improvements at each time point measure. All statistics are derived from two tailed paired t-tests based on comparisons between before and after of same color for each time point. *=p<0.05, **=p<0.01, ***=p<0.001, ****=p<0.0001. Error bars indicate estimated 95% confidence intervals.

### Age is not predictive of improvement in CRST scores

Previous findings have found that age plays little role in the effectiveness of MRgHIFU for treating ET (25). To assess whether age affects MRgHIFU effectiveness in our cohort, changes in CRST scores were assessed in individuals <70 years of age and ≥ 70 years of age. Age was not detected to affect MRgHIFU treatment effectiveness for ET in our analysis as there were no statistically significant differences in treatment effectiveness up to 3 months after treatment for either total CRST, treated hand specific Sub-A+Sub-B, or Sub-C scores (Figure 2). Therefore, our findings do not suggest that age negatively affects treatment effectiveness of MRgHIFU for ET.

**Figure 2.**
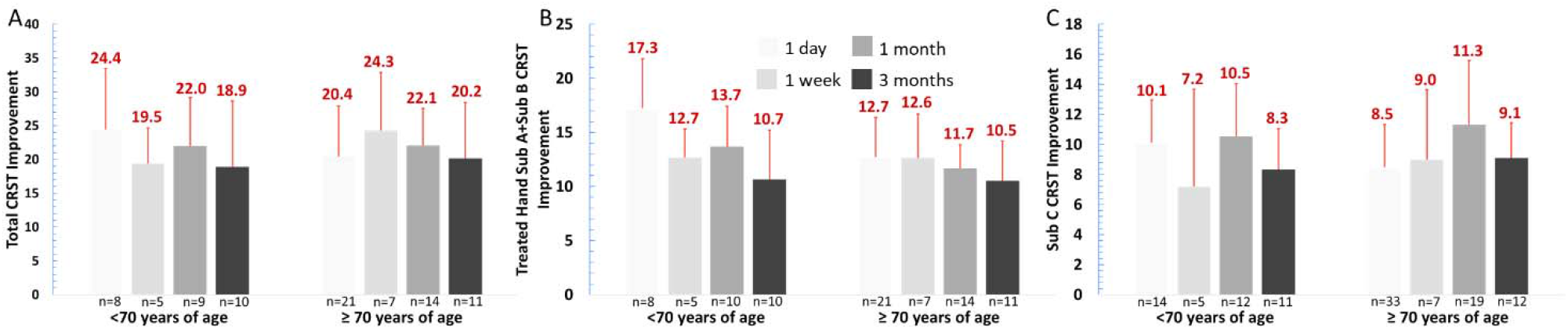
Total and CRST Subcategory improvements do not differ by age up to 3 months post HIFU. A: Total CRST score changes over time post-HIFU by age group. Total CRST score changes do not show a statistically significant difference by age group at each time measured post-HIFU. B: Treated Hand Sub A+Sub B score changes over time post-HIFU. More precise measurements of tremor severity changes in the affected upper extremity do not show statistically significant differences between age groups post-HIFU. C: Sub C CRST score improvements. Sub C score changes measuring functional disability related to ET improvement do not show statistically significant differences over time by age group. All statistics are derived from two tailed independent means t-tests with equal variances assumed based on comparisons between before and after of same color for each time point. Error bars indicate estimated 95% confidence intervals.

### Effects of T2D and CNCs on CRST scores

The role of comorbidities in predicting MRgHIFU treatment outcomes is not well understood. Therefore, several factors that could possibly affect outcomes were assessed including CVD, T2D, HTN, CNCs, pulmonary disease, kidney disease, liver disease, or previous stroke were assessed. Due to low sample size, our ability to assess the impact of liver disease or specific CNCs on tremor severity outcomes was limited. For all factors tested, only T2D and CNC were detected to affect improvement of CRST scores at different time points. T2D was detected as favorably affecting Sub C improvement at 1 month (Figure 3A). However, by 3 months, there was statistically significant impact thereby suggesting that the effects we detected are only transitory and not relevant to long term outcomes. Additionally, both those w/ and w/o T2D were detected to have statistically significant improvement from baseline to 1 month and the baselines between those w/ and w/o T2D were not found to be statistically different (Figure 3B). However, those w/ T2D were detected to have a lower Sub C CRST score at 1 month which may potentially indicate a more favorable outcome at this point in those w/ T2D (Figure 3B).

**Figure 3.**
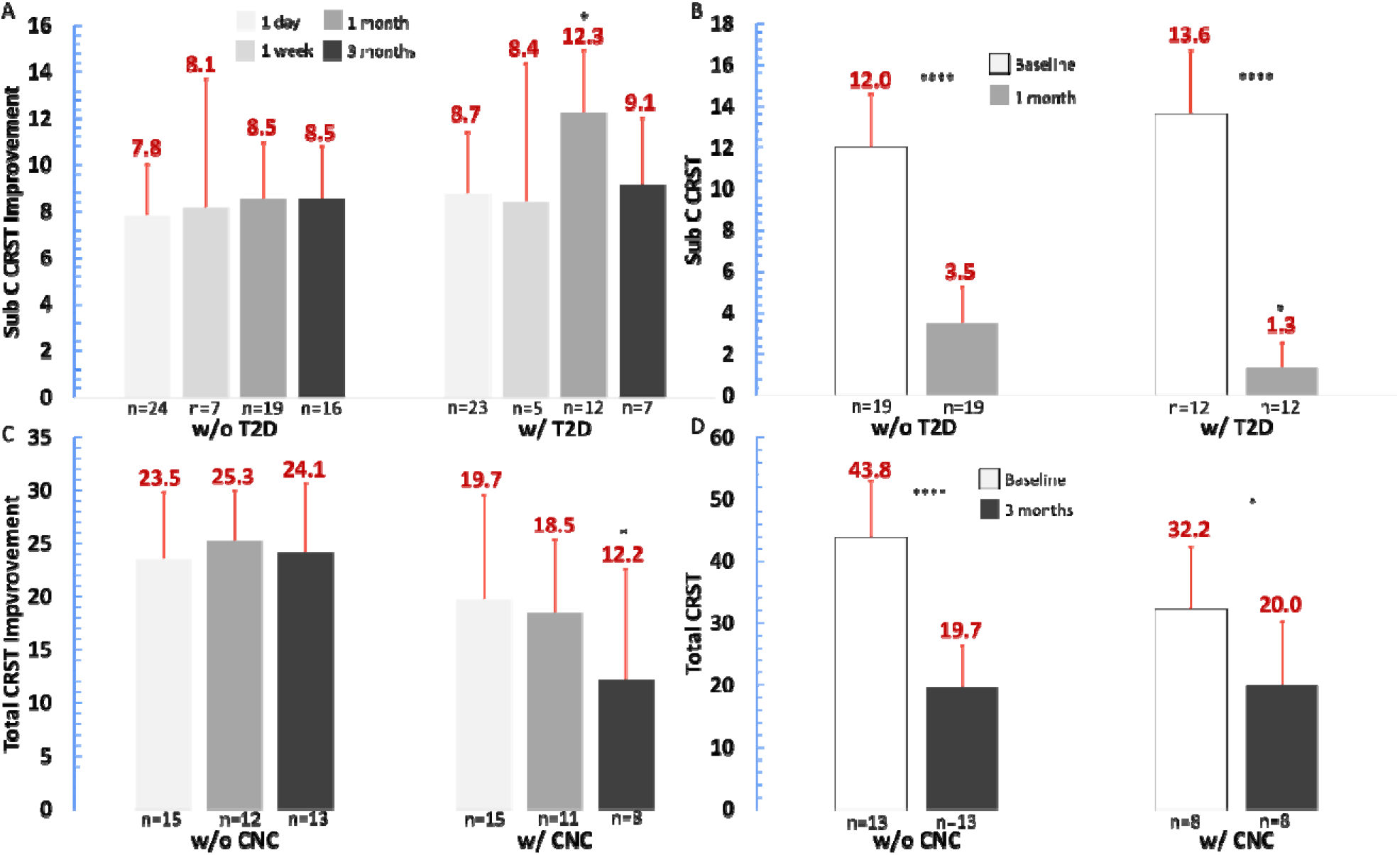
Sub C score among individuals with T2D and total CRST scores and among those with concurrent neurological condition (CNC). A: Sub C score improvements based on T2D status. measuring functional disability related to essential tremor showed statistically significant difference at 1 month post HIFU between those w/ and w/o T2D. B. Baseline vs 1 month Sub C CRST scores based on T2D status. Statistically significant differences found between baseline and 1 month for those both w/ and w/o T2D. However, 1 month time point also showed statistically significant difference between those w/ and w/o T2D as indicated by the asterisks above the 1-month time point in the w/ T2D columns. C. Total CRST scores improvements based on CNC presence. There is a statistically significant difference in total CRST score improvement at 3 months post HIFU between those w/ and w/o CNC. Note that the most common CNCs was Parkinsons’s disease (n=4). D: Total CRST scores between whose w/ and w/o CNC at baseline and 3 months. Baseline scores for both those w/ and w/o CNC were statistically different from the 3 month total CRST score for each group. However, there were no statistical differences between the baselines of each group or the 3 month time points of each group. Statistics in A and C derived from two tailed independent means t-tests with equal variances assumed for T2D comparison and CNC comparison based on comparisons between changes of same color denoted group for each time point. Paired two tailed t test used in B and D for differences between time points of same groups and two tailed independent means t-tests assuming equal variances used to compare baseline scores and 3 month time point scores based on T2D or CNC status for B and D *=p<0.05. Error bars indicate estimated 95% confidence intervals. *=p<0.05, **=p<0.01, ***=p<0.001, ****=p<0.0001.

CNCs were detected as having a statistically significant adverse impact on total CRST score improvements at 3 months (Figure 3C). However, further analysis was not found to detect a statistical difference in total CRST score outcome at 3 months and the baselines, while not statistically significant from each other, were higher for those w/o CNC (Figure 3D). Therefore, the apparently lower levels of improvement may be due to a higher baseline for those w/o CNC and outcomes were statistically the same at 3 months. The most common CNC in the 3-month cohort assessed was Parkinson’s disease (PD) (n=4). The relatively low number of patients at this time point for comparison limit further analysis into which specific CNC is likely causing the change.

### MRgHIFU is effective at improving QUEST scores up to 1 year post HIFU and age or comorbidities do not affect long term efficacy

MRgHIFU has been previously shown to improve QUEST scores that subjectively assess how a patient’s quality of life is affected by their tremor and our findings support this (Figure 4A). However, it is not clear whether age or comorbidities play a role in the efficacy of MRgHIFU in improving QUEST scores. Therefore, QUEST score changes were assessed at different time points up to 3 months and individuals <70 years of age or ≥70 years of age were not found to be statistically different (Figure 4B). Additionally, only CVD was found to affect QUEST score improvement, and this was only at 1 week post-HIFU with no statistical differences seen thereafter up to 3 months (Figure 4C). Additionally, the overall outcome of QUEST score at 1 week was not affected and both those w/ and w/o CVD showed improvements from baseline to 1 week (Figure 4D). Therefore, neither age nor the comorbidities nor possible confounding factors (family history of tremor and CNC) were found to negatively affect QUEST score improvements by 3 months.

**Figure 4.**
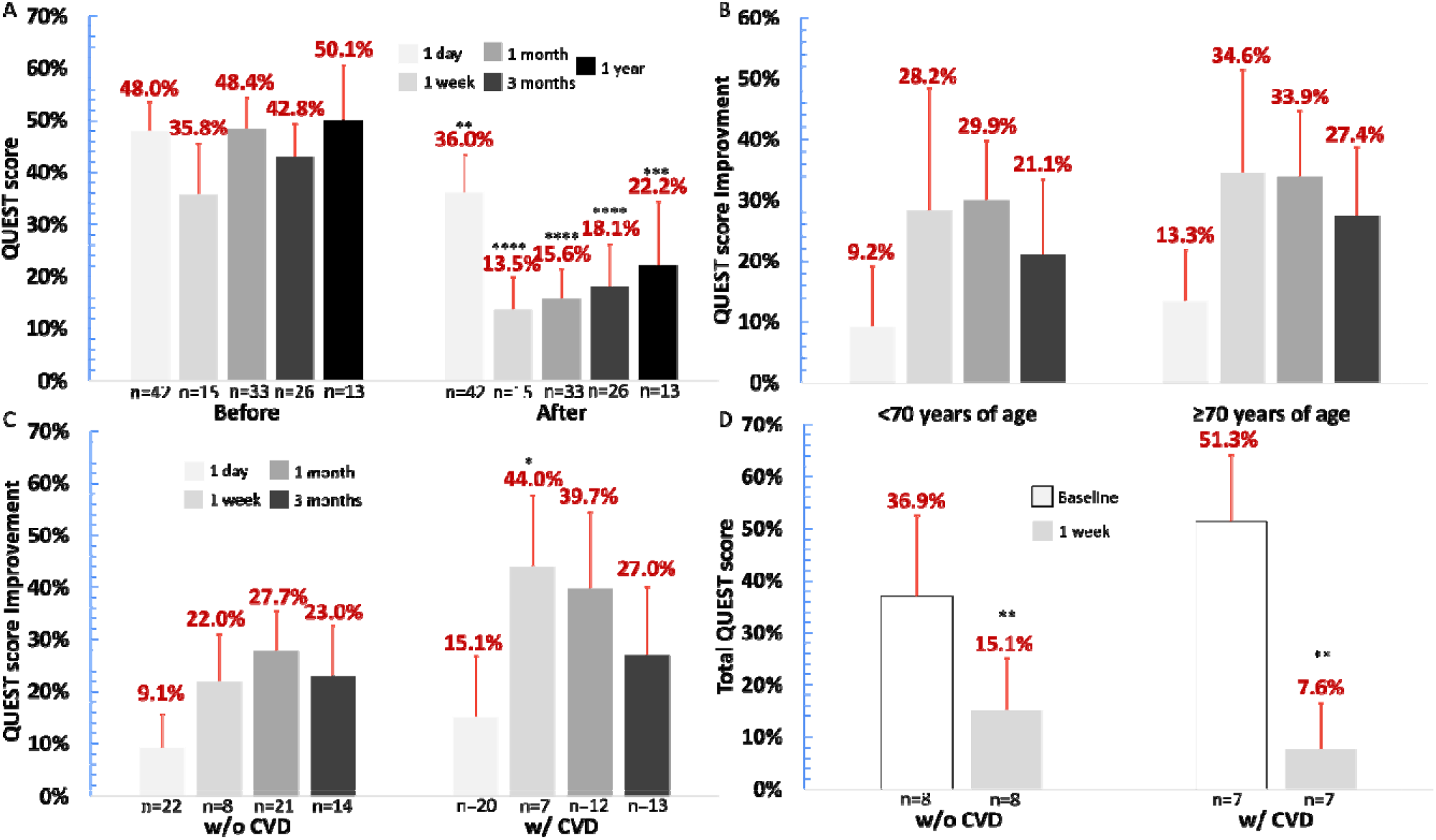
QUEST scores improve up to 1 year post HIFU and differences up to 3 months do not differ by age group or CVD status. A: QUEST scores up to 1 year post-HIFU. QUEST scores show a statistically significant decrease at all time points measured up to 1 year after HIFU. B: QUEST improvement changes by age group. No statistically significant differences in QUEST score improvement at any time point measured post-HIFU up to 3 months were found between age groups. C: QUEST improvements by CVD status. Individuals w/ CVD did experience more improvements in QUEST scores post HIFU at 1 week but this did not persist to the end of the time points measured at 3 months. D. Baseline vs 1 week post HIFU by CVD status. No differences between total QUEST score at baseline for those w/ or w/o CVD or at 1 week for those w/ or w/o CVD. However, the baseline QUEST score was statistically different from the 1 week QUEST score for both groups suggesting statistically significant improvement from baseline in each group as indicated by asterisks between the time points for each group. Paired two tailed t test used in A and D for differences between time points of same groups and two tailed independent means t-tests assuming equal variances used to compare baseline scores and 1 week time point scores based on CVD status for D. Differences in QUEST score improvements in both B and C assessed by two tailed independent means t tests with equal variances assumed. Error bars indicate estimated 95% confidence intervals. *=p<0.05, **=p<0.01, ***=p<0.001, ****=p<0.0001.

### Older adults do not show an increased side effect profile at any time point measured and comorbidities do not affect side effect profile long term

Age has been previously found to not play a role in the number of symptoms experienced by individuals treated with MRgHIFU for ET (26). However, only CVD and other vascular patholgies have been investigated as possible contributing factors to MRgHIFU efficacy (26).

Therefore, both possible confounding factors and comorbidities previously discussed were assessed at different points post-HIFU. Our cohort showed an array of side effects at all time points up to 1 year with the highest percentage of patients showing gait-balance changes (Table 2). We found that older adults experience fewer symptoms at 3 months compared to those <70 years of age (Figure 5A). Additionally, the data suggests that family history of tremor and pulmonary disease both transiently affect the number of symptoms experienced post-HIFU but this difference is no longer present at 3 months (Figure 5B-C). Overall, we did detect age or comorbidities as negatively affecting safety of MRgHIFU for ET. In fact, older adults report fewer side effects at 3 months. However, this finding may be the result of a type 1 error given the number of comparisons performed.

**Figure 5.**
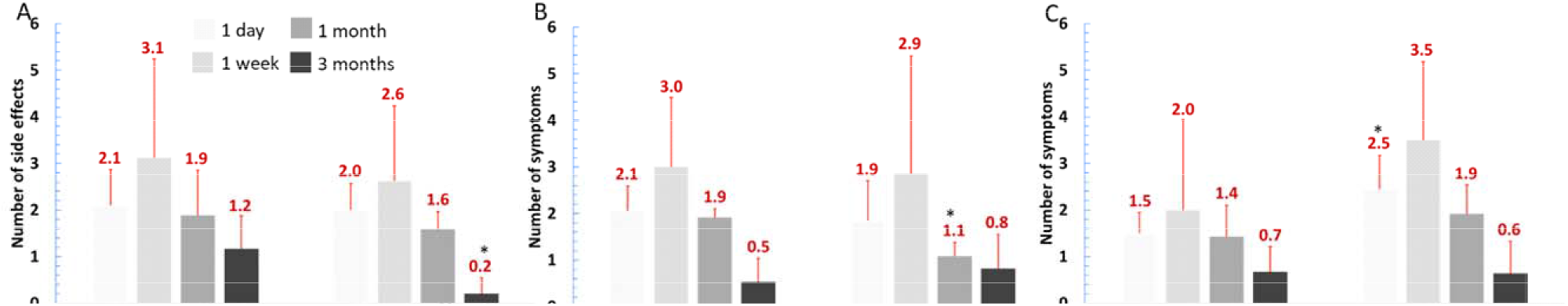
Symptoms following HIFU based on age, pulmonary disease status, and family history (Fam Hx) of ET. A: Symptoms are similar between age groups except for at 3 months where there is a statistically significant difference between the two groups. B: Symptoms based on pulmonary disease status are only statistically different at 1 month post HIFU with no statistical differences thereafter. C: Symptoms based on Fam Hx of ET are only statistically different at 1 day post HIFU with no statistical differences at later time points. All statistics derived from two tailed independent means t-tests with equal variances assumed for all not statistically significant comparisons. *=p<0.05. Error bars indicate estimated 95% confidence intervals.

**Table 2.**
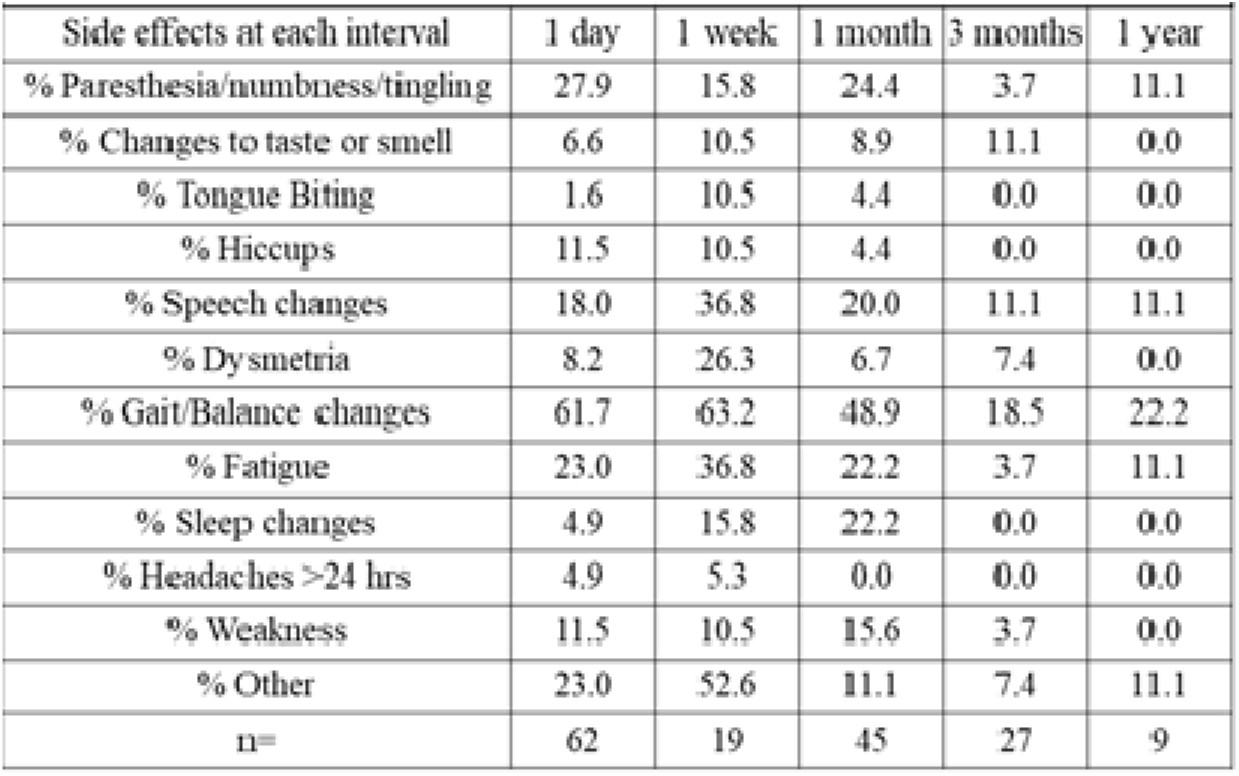
Data collected from EPIC records for patients at different time points show side effects over time up to 1-year post-HIFU. Note that gait/balance changes were the most common concerns over time. The most common “other” side effects at 1 day were vision changes (n=4) and dizziness (n=3), 1 week were drooling (n=3) and dizziness (n=2), and 1 month was vision changes (n=3).

## Discussion

Magnetic Resonance-guided High-Intensity Focused Ultrasound (MRgHIFU) thalamotomy is a well-established intervention for medication-refractory ET with a robust body of literature confirming its efficacy in reducing tremor severity and improving quality of life for up to four and five years (24–25, 27). While the procedure’s general effectiveness is well established, it is not fully understood how patient-specific factors, particularly advanced age and common medical comorbidities, influence long-term outcomes and safety. Our study directly addresses this gap, and our results did not detect that either age or a variety of comorbid conditions adversely affect the long-term efficacy or safety of MRgHIFU. Patients aged 70 years and older, as well as those with conditions such as cardiovascular disease, type 2 diabetes, and pulmonary disease achieved statistically similar improvements in tremor control, functional disability, and quality of life that were similar to those of younger and patients without the same comorbidities. Differential improvements at specific time points were noted for specific conditions but these findings should be applied to appropriate context. First, the main goal of MRgHIFU therapy is to improve ET in a targeted manner as measured by CRST Sub A+Sub B specific to the treated side and improve quality of life while maintaining safety. These end points were not adversely affected by any condition or age tested. Second, the low sample size at specific time points could introduce a higher level of variance with more room for influence and these differences may dissipate with more normal distributions from larger samples. Finally, the overall levels of CRST or QUEST scores were not statistically different at specific time points that showed statistical differences in improvements suggesting different baseline levels between groups, while not statistically detected, are likely the cause. Therefore, the evidence presented here did not detect that age and the comorbidities tested do not have an overall impact in outcomes of CRST or QUEST scores post-HIFU. Additionally, our research offers a significant and novel contribution for further exploration and validation by expanding beyond prior studies that focused primarily on age or vascular health alone (26).

A new observation from our data was that older patients (≥70 years) reported fewer total side effects at the 3-month follow-up compared to their younger counterparts. This finding, while requiring validation in larger cohorts, suggests the minimally invasive nature of MRgHIFU may be an advantage for older adults. Although counterintuitive, there are possible hypotheses that may explain this favorable safety profile and warrant future investigation. Alternatively, older patients may attribute side effects to normal aging rather than due to the procedure. Regardless, this finding did not support that age should be a deterrent in considering MRgHIFU for treatment of ET as the safety profile does not appear adversely affected by age as was also previously shown by others (26). Additionally, pulmonary disease and Fam Hx of tremor only had transient effects on side effects reported with no statistical differences found by 3 months.

The patients in our cohort that had both ET and PD meet the criteria for being categorized as ET-PD (28). Our study was not adequately powered to detect differences in efficacy between these two specific groups after finding that those with CNCs showed a statistically significant difference in total CRST change without affecting the total CRST endpoint. Additionally, UPDRS scores, rather than CRST scores are more appropriate to capture efficacy for improvement in PD. Future research should further examine efficacy for tremor treatment in this cohort using both CRST and UPDRS scores to improve our understanding of treatment responses in this population compared to ET or PD alone treatment.

From a clinical standpoint, our findings may have practical implications for patient selection and counseling. They support an understanding that comorbidity-related effects are likely transient and do not affect long term benefits from MRgHIFU treatment for ET. This is very important given the significant economic burden and healthcare costs associated with managing ET, which are driven in part by its high comorbidity burden (29). The consistent improvement we observed in both objective clinician assessments and subjective patient reports suggests that a broad spectrum of patients benefit similarly from MRgHIFU treatment for ET.

This study has limitations, including its single-center retrospective design and reliance on electronic medical record data for side effects, which may not capture all patient-reported experiences. Also, the electronic medical record often had incomplete and inconsistent follow up thereby leading to insufficient sample size available at 1 year for measuring how comorbidities may affect outcomes regarding CRST, QUEST, and side effects. While our data does not suggest there would be an effect, cohort studies with larger sample sizes are necessary to confirm this for longer term safety and efficacy. Furthermore, our patient population was predominantly Caucasian and therefore may not reflect whether specific differences in efficacy may occur for individuals of other ethnicities. Finally, statistics in this project were limited to excel within institutional constraints. Formal normality testing was not performed and formal correction for multiple comparisons was not applied thereby increasing the chance of false positive findings. Therefore, these findings should be regarded as exploratory and hypothesis generating in need of larger cohort confirmation and more robust statistical analysis.

In conclusion, this study suggests a durable efficacy of MRgHIFU treatment for ET and makes a significant novel contribution by suggesting that its benefits extend equally to older patients and those with a variety of common comorbidities and this occurs without a concomitant increase in long-term side effects. These results provide evidence to support the treatment of a more medically complex patient population which would thereby support the use of MRgHIFU for the treatment of ET across broad patient populations with a variety of comorbidities and into older age.

## Acknowledgements

The authors thank the Parkview Health Neuroscience Service Line, and Indiana University School of Medicine (IUSM) – Fort Wayne (FW) Student Education and Research Fellowship (SERF) program.

## Statements and Declarations

## Ethics Approval

This study was approved by the Institutional Review Board (IRB) of Parkview Health (IRB#: NEURO24-0412 HIFU). Because this was a chart review study, it was not feasible to obtain consent for data collection. Hence, the researchers successfully submitted and obtained Request for Waiver or Alteration of Consent and HIPAA authorization. All procedures performed were in accordance with the ethical standards of the institutional research committee and with the 1964 Helsinki Declaration and its later amendments.

## Consent to Publish

Not applicable as the manuscript does not contain any individual level participant data in any form (such as individual details, images, or videos).

## Data Availability

The data that support the findings of this study are available from the corresponding author upon reasonable request. The data are not publicly available due to privacy or ethical restrictions.

## Author Contributions

Adam Farmer: Data collection, conceptualization, statistical analysis, and writing of draft. William Young: Data collection, reviewing, and editing. Jody Neer: Supervision, reviewing, and editing. Fen-Lei Chang: Conceptualization, supervision, and data collection. Tara Kimbason: Conceptualization, data collection, and editing. All authors read and approved of the final manuscript draft.

